# Detection of *M. tuberculosis* in the environment as a tool for identifying high-risk locations for tuberculosis transmission

**DOI:** 10.1101/2022.04.27.22274350

**Authors:** Renu Verma, Flora Martinez Figueira Moreira, Agne Oliveira do Prado Morais, Katharine S. Walter, Paulo César Pereira dos Santos, Eugene Kim, Thiego Ramon Soares, Rafaele Carla Pivetta de Araujo, Bruna Oliveira da Silva, Andrea da Silva Santos, Julio Croda, Jason R. Andrews

## Abstract

Tuberculosis (TB) remains a leading cause of infectious mortality globally, yet most cases cannot be epidemiologically linked even with extensive contact investigations and whole genome sequencing. Consequently, there remain major gaps in our understanding of where and when *M. tuberculosis* (*Mtb*) exposures occur. We aimed to investigate whether *Mtb* can be detected in environments where TB patients were recently present, which could serve as a tool for characterizing exposure risk. We collected 389 environment surface (ES) swabs from two high TB burden prisons in Brazil, sampling 41 (n=340) cells occupied by individuals with active TB and 7 (n=49) cells from individuals without TB. In a subset of pooled swabs (n=6) and a swab from a cigarette lighter from the cell with active TB patients, we enriched *Mtb* DNA using RNA-bait hybrid capture assays and performed whole genome sequencing. In prison cells, *Mtb* DNA was detected in 55/340 (16%) of ES swabs from cells occupied by active TB patients and none (0/49) from cells in which no active TB patients were present. *Mtb* was detected in 13/16 (81%) prison cells occupied by the individuals with high/medium sputum Xpert *Mtb* load and 8/25 (32%) with low/very low sputum *Mtb* load (p=0.003). Seven hybrid capture samples had a median genomic coverage of 140X. *rpoB* mutations conferring high-level rifampin resistance were detected in 3/7 ES swabs. *Mtb* was frequently detectable in environments recently occupied by individuals with active TB. This approach could be applied in congregate environments to identify and characterize high-risk settings for *Mtb* exposure.

## 1. Introduction

Despite the availability of highly effective treatment regimens and reliable rapid diagnostics, more than 10 million new cases of TB are estimated to occur every year ^1^. A major obstacle to reducing this burden is ongoing transmission of infection. Classical TB transmission studies predominantly rely on detecting *Mtb* either from patient-derived samples such as sputum or bioaerosols generated by an active TB patient at a single time-point^2-4^. However, a major limitation of this approach is that infectious individuals remain undiagnosed for several weeks to months, making it challenging to identify locations of transmission^5^. Using molecular data combined with household and social contacts, studies have reported that only a minority of *Mtb* infections in adults are attributable to these identifiable contacts^6-8^. A study in KwaZulu-Natal in South Africa found that only 12% of the transmission was molecularly linked to close contacts^9^. In another HIV-prevalence community in South Africa, only 19% of clustered TB cases could be attributed to household transmission^10^. These challenges have also been independently observed in rural Malawi where only 9% of TB cases were attributable to known contacts^11^. Taken together, these findings suggest that most *Mtb* infections are acquired in congregate settings outside the household; however, there is little direct evidence about where such transmission occurs.

Several emerging approaches have been undertaken in attempts to identify high risk locations for *Mtb* exposure in community settings. These include social network analysis among individuals diagnosed with TB^12^, use of wearable GPS loggers to identify durations individuals spend in various environments^13^, and mapping of ‘rebreathed air’ across communal environments using portable carbon dioxide monitors^14^. These approaches provide insights into potential exposure risks, though such evidence is indirect in the absence of *Mtb* detection in these environments. We hypothesized that *Mtb* DNA could be detectable on surfaces in environments where active TB patients were recently present, providing direct evidence about exposure risk in these environments. To test this hypothesis, we developed a scalable environmental sampling and molecular testing approach for *Mtb* DNA detection and validated it in a very high-risk environment--prisons. We further demonstrated that *Mtb* whole genome sequences can be recovered from environmental samples using an RNA-bait hybrid capture approach.

## 2. Methods

### 2.1. Ethics statement

The clinical study was approved by the institutional review boards (IRB) of the Federal University of Grande Dourados, Brazil (IRB##3.483.377) and Stanford University (IRB# 40285). All participants were over 18 years of age and provided written informed consent to participate.

### 2.2. Tuberculosis screening in prisons

As a part of an active case finding program, our team performed mass screening for TB in two prisons in Brazil (Penitenciária Estadual de Dourados and Estabelecimento Penal Jair Ferreira de Carvalho) by chest radiography and sputum testing by GeneXpert MTB/RIF Ultra assay and culture^15^. All incarcerated individuals were offered screening and >90% agreed to participate. Sputum testing was performed for all individuals who were able to produce sputum, irrespective of reported symptoms. Based on these data, we identified 41 cells occupied by active TB patients, in which we undertook environmental surface sampling.

### 2.3. Assay development and validation

We first performed a pilot study at Stanford Hospital, CA, USA, to develop and validate the environment sampling assays (Supplemental methods and results). Sterile Hydraflock swabs (Longhorn Primeswab LH-11-10) were used to collect ES swabs. Surfaces covering an area roughly the size of an adult palm (approx. 70 cm^2^) were swabbed with a hyraflock swab that was pre-wet with buffer (1X TE or 1X PBS). After collection, the ES swabs were immediately dispensed in 500ul buffer and transported to the molecular lab. We tested the *Mtb* DNA detection methods on two different platforms: GeneXpert MTB/RIF Ultra (Cepheid, Sunnyvale, USA) and StepOnePlus real time PCR machine (Applied Biosystems) using insertion sequence (IS) 6110 and the region of difference 9 (RD-9) custom TaqMan assays described previsouly^16^. IS6110 is typically present in multiple, but variable numbers of copies in the *Mtb* genome, making it a sensitive target for detection but precluding quantification of genome copies, whereas RD-9 is present as a single copy, enabling genome copy quantification.

#### 2.3.1 Custom TaqMan assays

For custom TaqMan assays, we collected ES swabs in 500ul 1X TE buffer (pH=7.4) (Millipore Sigma) for DNA extraction. DNA was extracted using QIAamp DNA mini kit (Cat # 51304). All qPCR reactions were performed in 20ul volume using TaqMan™ Environmental Master Mix 2.0 (Cat# 4396838) with 500nM of each forward and reverse primers and 250nM of probes. The samples were analyzed in technical duplicates on a StepOnePlus real time PCR using the following cycling conditions; initial denaturation (95°C;10 minutes), 40 cycles (denaturation 95 °C 15 seconds and 60°C 30 seconds annealing and extension). The samples were considered positive only when the amplification was observed in both duplicates of IS6110 and RD-9 assays. DNA copies were quantified using a RD-9 standard curve derived from *Mtb* H37Rv ATCC strain (Catalogue#25618DQ) (**Supplementary Figure 1**).

#### 2.3.2 Analysis on GeneXpert MTB/RIF Ultra

For GeneXpert analysis, we collected ES swabs in 500ul sterile 1X PBS (pH=7.4) (Cytiva HyClone™) and used directly without manual DNA extraction. Briefly, the swabs were vortexed for 30 seconds and the samples were mixed with Xpert MTB/RIF buffer in 1:1 (sample to buffer ratio). The samples were vortexed for 5 seconds followed by 5 minutes incubation at room temperature. The entire 1ml sample was loaded into GeneXpert MTB/RIF Ultra cartridge for *Mtb* DNA detection. GeneXpert *Mtb* load was graded semiquantitatively as high (Ct <16), medium (Ct 16<22), low (Ct 22–28), very low (Ct >28) and trace).

### 2.4. Environmental surface sampling in prison cells

We collected a total 389 ES swabs from 48 cells. Forty-one cells were occupied by at least one individual with TB diagnosed within the past 14 days. Seven prison cells were occupied by individuals without known active TB (**Table 1**). We collected 340 swabs from cells with active TB and 49 swabs from cells without active TB patients. For security reasons, the ES swab samples were collected by the incarcerated individuals who were provided brief instructions by the study staff. Prior swab collection, study staff briefly explained the purpose and importance of collecting ES swabs from the prison cells followed by a demonstration on the swabbing technique and dispensing the swab in the collection tube. The study staff waited outside the cell instructing the incarcerated individuals about which surfaces they should swab **(Supplementary Figure 2)**. The swabs were collected in five categories-clothing, bedding, bathroom, personal utensils, and other surfaces in the cell which included floors, windows, walls, ceilings, and doors. As additional low-risk control sites, we collected 70 additional ES swabs from apartments (n=7) and a non-TB lab (n=1) at the Federal University of Grande Dourados, Brazil.

**Table 1.** Characteristics of individuals with active tuberculosis in study prison cells selected for environmental sampling.

| Sampling sites | Prison EPJFC | Prison PED | Total (n) |
| --- | --- | --- | --- |
| Number of cells sampled | 26 | 15 | 41 |
| Individuals with active TB | 28 | 17 | 45 |
| Individuals with cough | 13/28 (46 %) | 9/17 (53%) | 22/45 (49%) |
| **Bacillary load in sputum samples based on GeneXpert grading\*** |  |  |  |
| High | 6 (21%) | 8 (47%) | 14 (31%) |
| Medium | 5 (18%) | 1 (6%) | 6 (13%) |
| Low | 8 (29%) | 6 (35%) | 14 (31%) |
| Very low | 9 (32%) | 2 (12%) | 11 (24%) |

**Table 2.** Detection of *Mtb* DNA on environmental surface swabs from prison cells.

|  | ES swab sampling sites |  |  |
| --- | --- | --- | --- |
|  | Prison EPJFC | Prison PED | Total (n) |
| **Number of prison cells sampled** | 26 | 15 | 41 |
| **Surfaces swabbed** | 189 | 151 | 340 |
| **Swabs positive on GeneXpert** | 19/119 (16%) | 30/151 (20%) | 49/270 (18%) |
| **Swabs positive on StepOne Plus** | 6/70 (9%) | -- | 6/70 (9%) |
| **Total Positive** | 25/189 (13%) | 30/151 (20%) | 55/340 (16%) |

### 2.5. Sample collection for whole genome sequencing

Based on the GeneXpert results on ES swabs, we identified seven cells with highest rates of *Mtb* DNA positivity and collected additional 85 ES swabs for DNA enrichment with RNA baits-based hybrid capture assays and whole genome sequencing. The swabs were collected in 500ul 1X TE buffer. We first processed 15 swabs randomly chosen from a batch of 85 samples and individually analyzed those on IS6110 TaqMan assay. Only one swab collected from a cigarette lighter which was earlier positive on GeneXpert MTB/RIF Ultra was positive on the custom TaqMan assay. Therefore, to maximize the chances of detecting *Mtb* DNA, instead of analyzing the swabs individually, we divided the swabs from each cell into three categories-utensils, bed & clothing and other surfaces. We pooled the samples within each category before DNA elution to get total three swabs (one from each category) per cell. DNA was extracted as described above and eluted in 30ul nuclease-free water.

### 2.6. *Mtb* culture from paired sputum samples

To compare the enriched genomes of ES swab samples with the genomes of *Mtb* present in the sputum of individuals occupied those cells at the time of swab sampling, we further cultured *Mtb* from three sputum samples using Ogawa-Kudoh Method described previously^17^. Briefly, a sterile cotton swab was dipped and swirled in 2ml sputum and submerged into a 4% solution of sodium hydroxide (NaOH) for 2 minutes to decontaminate the sample. The excess of 4% NaOH was removed by compression of the swab against the flask wall and the sample was inoculated in Ogawa-Kudoh medium (pH 6.4). The cultures were incubated at 37°C and observed for growth twice a week for 60 days. *Mtb* DNA was extracted using Cetyltrimethyl ammonium bromide (CTAB) method described previously^16^.

### 2.7. Hybrid capture assays and whole genome sequencing

We performed IS6110 TaqMan qPCR on DNA extracted from 21 pooled swabs corresponding to seven cells. Six pooled samples from three cells and one swab from a cigarette lighter, all positive on IS6110 assay, were considered for hybrid capture assays. DNA from ATCC H37Rv was used as a positive control. All sequencing libraries were prepared using Illumina DNA Prep library preparation kit (Cat # 20018704) following manufacturer instructions. Following library preparation, we pooled eight libraries consisting of seven samples from prison cells and one with H37Rv DNA (ATCC; 10^4^ copies). To achieve maximum genome coverage in hybrid capture reactions, we split the libraries into 4 parts (7ul each) and enriched each library using biotinylated RNA-baits (myBaits®; Arbor Biosciences) based on the inferred ancestral genome of the *Mtb* complex, which is genetically equidistant to all *Mtb* lineages. We performed the baits protocol for four replicate reactions following the standard protocol published by the manufacturer using a hybridization and wash temperature of 60°C. Briefly, the concentrated library pools were first split into four reactions, each undergoing hybridization incubation (60°C, 24 hours). These reactions were then bound to streptavidin-coated magnetic beads and washed (60°C). Finally, each reaction was split into two 15uL aliquots and performed a post-capture PCR amplification of both aliquots (8 total PCR reactions) using 2x KAPA HiFi HotStart Ready Mix (Roche) and 10x KAPA Library Quantification Primer Mix (Roche). The post-capture amplification reaction consisted of an initial denaturation step at 98°C (2 minutes), then 12 cycles of 98°C (20 seconds), 60°C (30 seconds), and 72°C (45 seconds). Final extension was at 72°C (5 minutes) and held at 8°C until 0.8x Agencourt Ampure SPRI beads cleanup. A total 20uL of cleaned enrichments were analyzed on the Illumina Miseq in paired-end runs of 2×292bp length.

### 2.8. Whole genome sequencing and bioinformatic methods

We trimmed low-quality bases (Phred-scaled base quality < 20) and removed adapters with Trim Galore (stringency=3)^18^. We used CutAdapt to further filter reads (--nextseq-trim=20 -- minimum-length=20 --pair-filter=any) ^19^.To exclude potential contamination, we used Kraken2 to taxonomically classify reads and removed reads that were not assigned to the *Mycobacterium* genus or that were assigned to a *Mycobacterium* species other than *Mtb* ^20^. We mapped reads with bwa v. 0.7.15 (bwa mem)^21^ to the H37Rv reference genome (NCBI Accession: NC_000962.3) and removed duplicates with sambamba ^22^. We called variants with GATK 4.1 HaplotypeCaller^23^, setting sample ploidy to 1, and GenotypeGVCFs, including non-variant sites in output VCF files. We included variant sites with a minimum depth of 10X and a minimum variant quality score 40 and constructed consensus sequences with bcftools consensus^24^, excluding indels, and used SNP-sites to extract a multiple alignment of SNPs between sampled genomes^25^. We excluded SNPs in previously defined repetitive regions (PPE and PE-PGRS genes, phages, insertion sequences and repeats longer than 50 bp)^26^. We measured drug resistance associated mutations with MykrobePredictor v0.8.0, using the ‘201901’ database of genomic predictors of resistance^27^. We identified lineage and drug resistance associated mutations with TBProfiler v.2.8.6 ^28^. Our bioinformatic pipeline is available at https://github.com/ksw9/mtb_pipeline.

### 2.9. Phylogenetic analysis

We used the R package *ape* to measure the number of pairwise SNP differences between samples^21^. Missing sites, “Ns” in the alignment, were not counted as pairwise differences. To explore if heterozygous calls in samples with mixed infections were contributing to observed pairwise distances between samples, we excluded heterozygous sites with a minor allele frequency >5% using *bcftools* and recalculated pairwise SNP distances.

We fit maximum likelihood trees with RAxML-ng 1.0.1 and used a GTR substitution model and a Stamatakis ascertainment bias correction for invariant sites in our alignment. We divided the number of invariant sites by 1000 to avoid issues created by small branch lengths. We defined nucleotide stationary frequencies as frequencies in the reference genome.

### 2.10. Data analysis

We compared the detection of at least one swab in a cell by GeneXpert semi-quantitative grade using Fisher’s exact test. We used mixed effects logistic regression with random effect for cell to assess the relationship between Xpert semiquantitative grade and swab positivity. All statistical analyses were performed using R^29^.

## 3. Results

### 3.1. *Mtb* DNA detection in prison cells

We sampled 48 prison cells, 41 of which were occupied by 45 active TB patients. At any time point during swab collection, each cell was occupied with multiple individuals without TB sharing a cell with active TB patients (median= 14; IQR, 11-17). Thirty-seven cells housed one active TB patient each, whereas four cells accommodated two active patients. Approximately half (22/45; 49%) of participants with diagnosed TB reported cough at the time of swab collection. At the time of diagnosis, sputum GeneXpert semi-quantitative grade was high in 14/45 (31%) participants, medium in 6/45 (13%), low in 14/45 (31%), and very low in 11/45 (24%). TB treatment had already been initiated for 56% (25/45) for a median of 3 days (IQR, 1-10) at the time of sample collection (**Table 1**).

Of the 340 swabs collected from 41 prison cells with active TB patients, *Mtb* DNA was detected in 55/340 (16%) of ES swabs. We did not detect any *Mtb* DNA (0/49) in swabs collected from non-TB lab and residential apartments which served as negative controls. *Mtb* DNA positivity on custom TaqMan assays was 6/70 (9%) which was relatively higher on GeneXpert MTB/RIF Ultra 49/270 (18%). Among those positive on GeneXpert, the majority were trace (37/49; 76%) and remainder were low (5/49; 10%) or very low (7/49; 14%). *Mtb* DNA was detected on various surfaces, and positivity did not significantly differ across surface location or type: clothing, 25% (2/8); bedding, 23% (16/71); bathroom surfaces, 17% (8/48); personal utensils, 14% (6/42); other surfaces in the cell, 13% (23/171) (**Figure 1A**).

**Figure 1.**
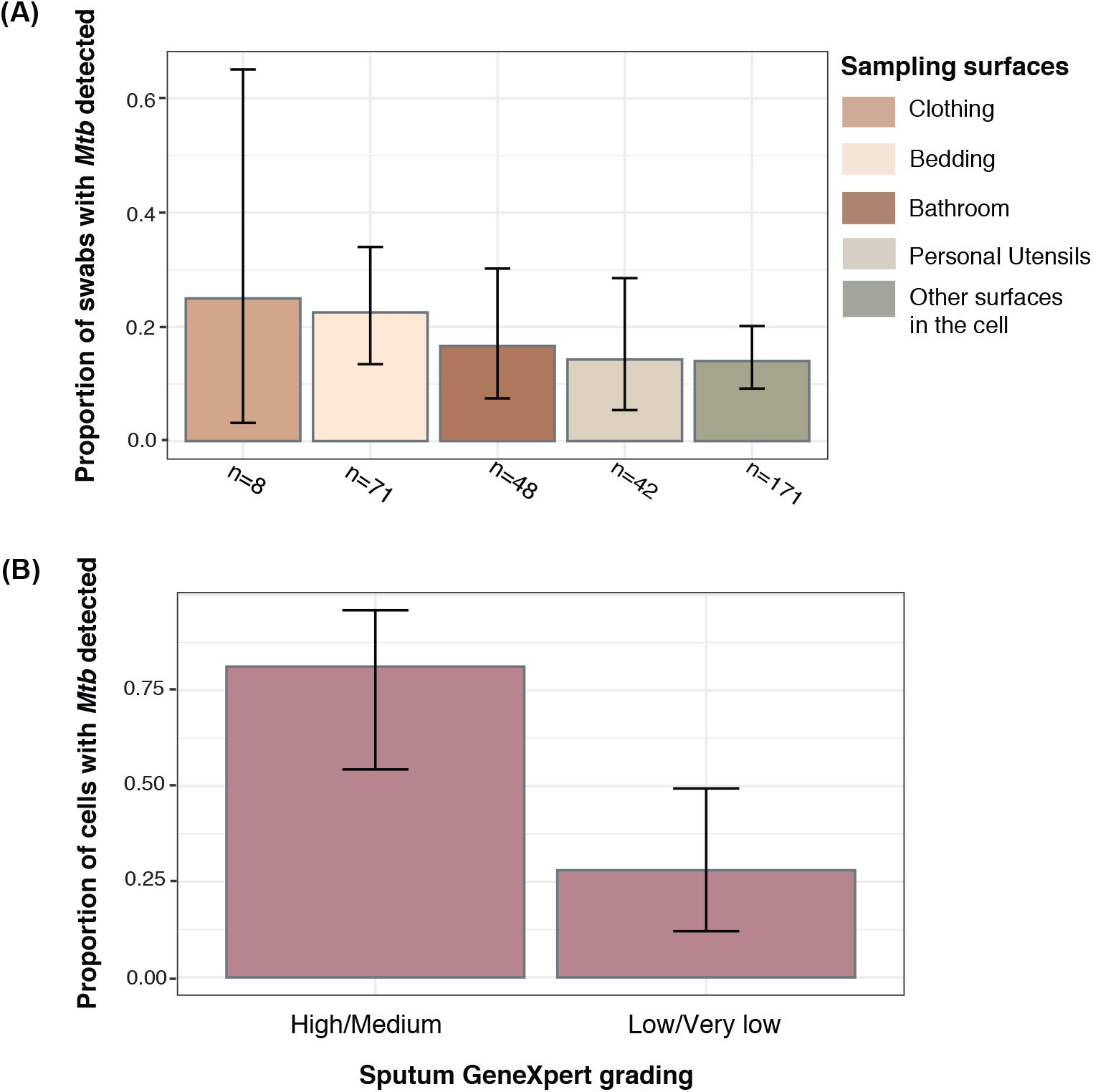
(A) *Mtb* PCR positivity by surface type in prison cells. The samples were divided into five different categories- clothing (towel, shirt, trousers); bedding (bedsheets, pillow covers, blanket and mattress); bathroom (sink, faucet, shower handle); personal utensils (cup, bowl, spoon and cigarette lighter); other surfaces in the cell (floor, wall, window, ceiling). **(B) Proportion of cells with positive *Mtb* PCR on ES swab**. The cells were classified into two groups (high/medium and low/very low) based on sputum Xpert bacillary load of the active TB patient present in the cell at the time of swab collection. The cell was counted as positive if *Mtb* DNA was detected in at least one swab on GeneXpert MTB/RIF Ultra or RD-9 TaqMan assay.

**Figure 2.**
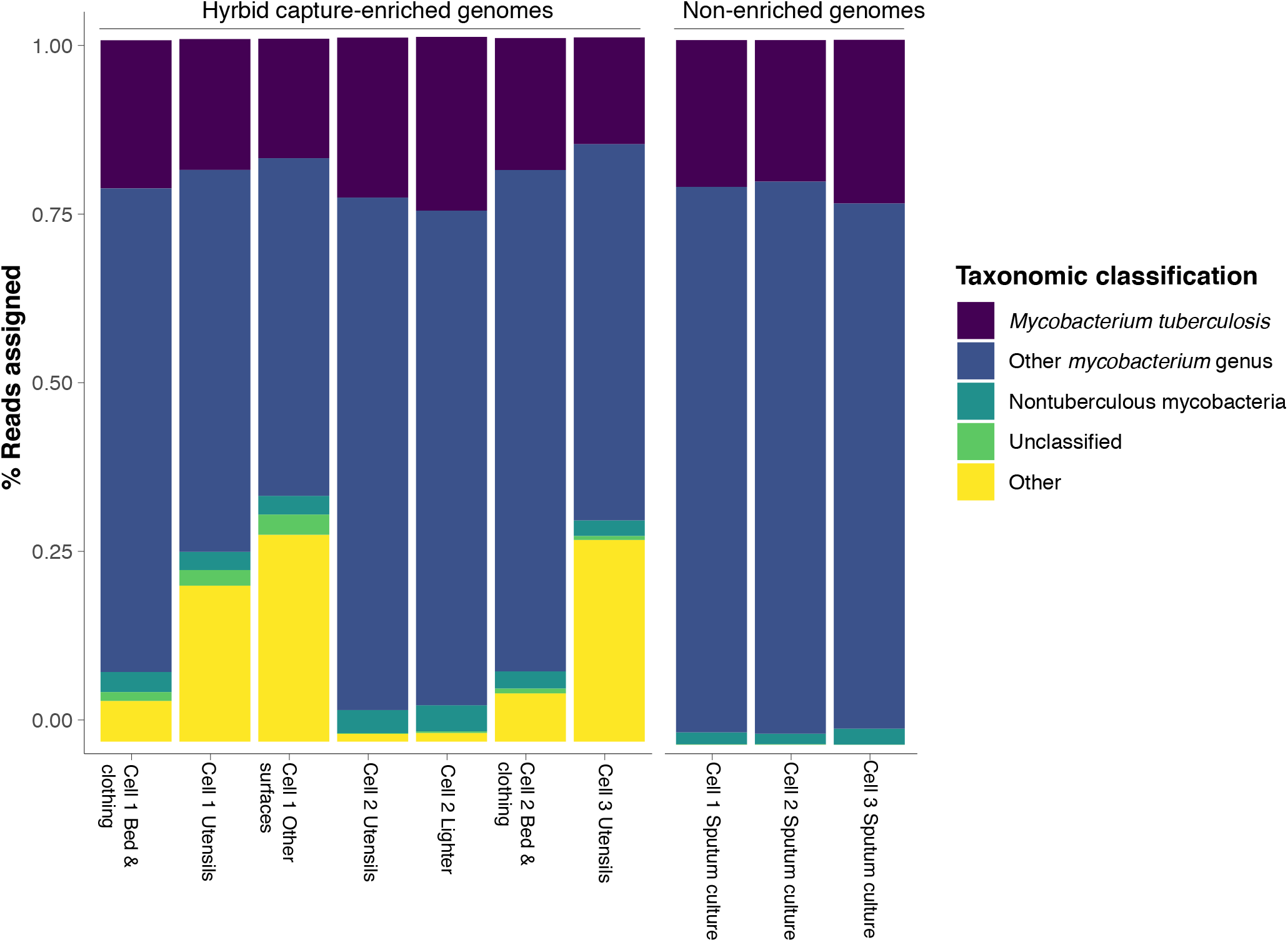
Proportions of reads taxonomically assigned to the *Mycobacterium tuberculosis* species complex, the *Mycobacterium* genus (excluding the *Mtb* species complex or nontuberculous *Mycobacteria)*, nontuberculous *Mycobacteria*, unclassified, or mapping to other genera. Facets indicate the type of sample source: hybrid capture enriched genomes from environment swabs and non-enriched genomes from sputum culture. Taxonomic classification was performed on trimmed reads using Kraken2.

Of the 41 cells occupied by active TB patients, *Mtb* was detected in at least one sample in 13/16 (81%) of the cells housing TB patients with high or medium Xpert sputum bacillary load and in 8/25 (32%) of the cells occupied by individuals with low or very low sputum *Mtb* load (p=0.003; **Figure 1B**). On a per-sample basis, 27% of samples (44/161) were positive from cells with medium or high sputum Xpert cases and 6% of samples (11/179) were positive from cells with low or very low sputum Xpert. *Mtb* detection on ES swabs was positively correlated with sputum Xpert bacillary load (p<0.001)

### 3.2. Hybrid capture assays and whole genome sequencing

Because of the relatively low quantity of *Mtb* DNA in ES swab samples compared to routinely sequenced cultured sputum, we took an RNA hybrid capture approach, allowing us to selectively enrich for *Mtb* DNA from a mixed template. Six pooled swabs including three from utensils, two from bed and clothing, and 1 from other surfaces in addition to one swab from a cigarette lighter corresponding to three cells were positive on IS6110 qPCR and were selected for hybrid capture and whole genome sequencing (**Table 3**). To compare captured ES swab samples to sequenced cultured sputum, we additionally sequenced three cultured sputum samples from the *Mtb* positive individual in the same cells.

**Table 3.**
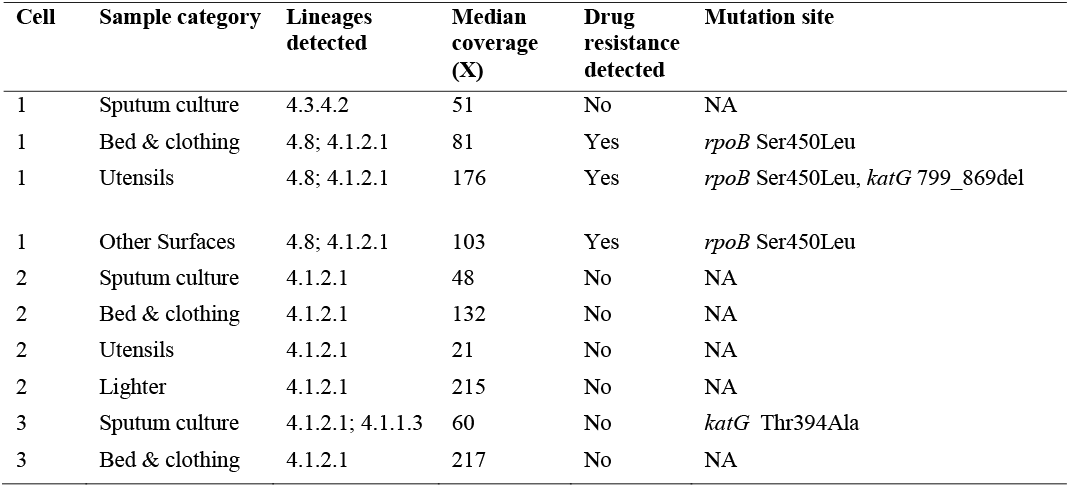
Hybrid capture assay and sequencing statistics for environment samples and samples extracted from matched sputum cultures.

| Cell | Sample category | Lineages detected | Median coverage (X) | Drug resistance detected | Mutation site |
| --- | --- | --- | --- | --- | --- |
| 1 | Sputum culture | 4.3.4.2 | 51 | No | NA |
| 1 | Bed & clothing | 4.8; 4.1.2.1 | 81 | Yes | *rpoB* Ser450Leu |
| 1 | Utensils | 4.8; 4.1.2.1 | 176 | Yes | *rpoB* Ser450Leu, *katG* 799_869del |
| 1 | Other Surfaces | 4.8; 4.1.2.1 | 103 | Yes | *rpoB* Ser450Leu |
| 2 | Sputum culture | 4.1.2.1 | 48 | No | NA |
| 2 | Bed & clothing | 4.1.2.1 | 132 | No | NA |
| 2 | Utensils | 4.1.2.1 | 21 | No | NA |
| 2 | Lighter | 4.1.2.1 | 215 | No | NA |
| 3 | Sputum culture | 4.1.2.1; 4.1.1.3 | 60 | No | *katG* Thr394Ala |
| 3 | Bed & clothing | 4.1.2.1 | 217 | No | NA |

The seven captured ES swab samples had a median genomic coverage of 140X (range 23 to 209) (**Supplementary Figure 2**). A mean of 98% of the *Mtb* genome had a depth of at least 10X; 65% had a depth of at least 100X. The median efficiency of our hybrid capture assay, which we defined as the percentage of total sequenced reads that taxonomically assigned to the *Mycobacterium* genus and were not assigned to a nontuberculous *Mycobacterium* species was 92% (range 65% to 98%).

### 3.3. Phylogenetic analysis

*Mtb* sequence data from captured ES swabs allowed us to assign taxonomy and investigate phylogenetic relatedness between samples. All seven captured ES swab samples were assigned to lineage 4, the predominant lineage in the state, and nested within previously sampled genomic diversity from the state^30^ (**Figure 3**).

**Figure 3.**
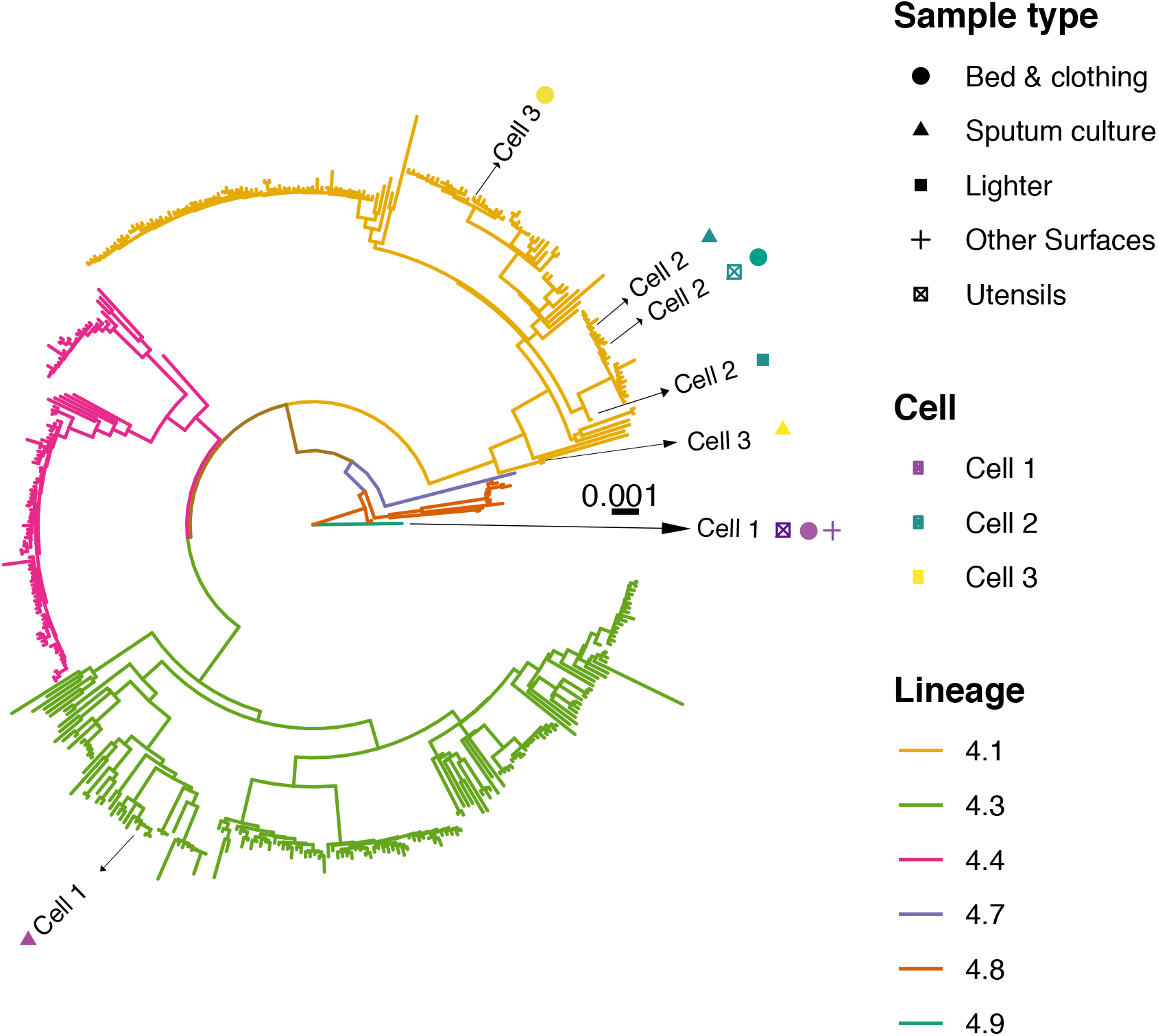
Maximum likelihood tree of 438 *Mtb* isolates from Mato Grosso do Sul, Brazil. The hybrid captured environmental and culture samples from patients within corresponding cells nest within circulating *Mtb* diversity sampled through active and passive surveillance in the state. For the paired environmental and culture samples, tip point colors and labels indicate cell and tip shapes indicate sample type. Phylogenetic branches are colored by assigned sublineage. Branch lengths are in units of substitution per site.

The three ES swabs from cell 1 were closely related (pairwise distance: 0-2 SNPs). In all three, we found evidence that both sub-lineages 4.8 and 4.1.2.1 were present; while the culture from that cell was assigned to lineage 4.3.4.2 (Table 3). We predicted that consensus sequences generated from environmental samples with two distinct sub-lineages could be chimeric, thus overestimating the genetic distance between these consensus sequences and the matched culture. To test this, we excluded heterozygous sites (Methods) from the environmental sample consensus sequences. Revised consensus sequences from the three ES swab samples were identical to one another and were 23 SNPs distant from the matched cultured isolate in cell 1 (85 SNPs distant before excluding heterozygous sites).

The three captured ES swab samples and one cultured sample from Cell 2 were all assigned to sub-lineage 4.1.2.1 and were closely related, with consensus sequences 0-3 SNPs distant (Table 3). When excluding heterozygous sites (Methods), consensus sequences were 0-2 SNPs distant.

Both sub-lineages 4.1.2.1 and 4.1.1.3 were present in the single captured ES swab sample from cell 3 and the matched cultured sample was assigned to sub-lineage 4.1.2.1 (Table 3). Again, we predicted that the presence of multiple strains in the could lead to an over-estimate of genetic distances between samples. We found that the environmental samples were genetically distant from the cultured isolate from the same cell (pairwise distance: 55 SNPs; when excluding heterozygous sites: 23 SNPs).

### 3.4. Antibiotic resistance prediction

We predicted antimicrobial resistance profiles using a database of confirmed mutations associated with resistance phenotypes^31^. All three of the captured ES swab samples from cell 1 carried the same rifampicin resistance conferring mutation (*rpoB* Ser450Leu). All three of these environmental samples were heteroresistant: both the susceptible and resistant alleles were present at moderate coverage depth, possibly on the distinct sub-lineages identified. 71% (50/70) of reads corresponded to the resistance-conferring mutation for the bed & clothing sample, 63% (85/134) for the utensils sample, and 60% (47/78) from the other surfaces sample. Additionally, a 70 base pairs deletion (799-869) in the *katG* gene associated with isoniazid resistance, was present in 18% of reads in the cell 1 utensils sample. It is likely that the *rpoB* and *katG* mutations, supported by 63% and 18% of reads respectively, were on different genomic backgrounds and do not reflect the presence of multi-drug resistant *Mtb*. The corresponding culture sample from a patient in cell 1 was drug sensitive. The remaining captured ES swab samples and corresponding cultures from the two other cells contained no known drug-resistance conferring mutations.

## 4. Discussion

In this study, we report the development and testing of a novel sampling approach based on detecting *Mtb* DNA on environment surfaces where active TB patients were present. We found that *Mtb* DNA was detectable in most environments where individuals with active TB had recently resided, and that detection was positively correlated with sputum bacillary load. By adapting our sampling method for GeneXpert, which is widely available in tuberculosis-endemic countries, this approach could be utilized in resource-constrained settings with minimal training or equipment. We further found that we could sequence *Mtb* genomes directly from environmental samples by using an RNA-bait hybrid capture approach, achieving sufficient coverage for lineage identification, phylogenetic analyses, and variant calling for resistance-associated mutations. Taken together, these findings suggest that environmental sampling could be a feasible approach for generating new dimensions of data for studying tuberculosis transmission.

Several historical studies in late 19^th^ and early 20th century demonstrated the detection of *Mtb* in natural and built environments. In 1888, Cornet collected dust from TB medical wards, residential apartments, and asylums with active TB patients. The dust was subcutaneously inoculated into guinea pigs. Among those inoculated with dust from TB sites, 16.4% developed TB, while none of the guinea pigs in negative control group developed TB^32^. Later, in 1920, Rogers conducted similar experiments by treating dust with 2% sodium hydroxide solution subsequently inoculating the guinea pigs. In a series of experiments, seven out of ten guinea pigs were diagnosed with disseminated TB^33^. While these findings provide compelling evidence that *Mtb* may be detectable in the environment, most of these studies were performed in an era when identification of *Mtb* at molecular level was not available. This is particularly important in the context of environment samples, where slow growing *Mtb* can be outgrown by other mycobacteria, which may also share similar clinical or phenotypic features. In 2015, Velayati et al, attempted to isolate *Mtb* from 1,500 randomly collected soil and water samples from three counties in Tehran^34^. The samples were inoculated on Löwenstein–Jensen culture media and monitored for growth for several weeks. *Mtb* growth was observed in 1% soil and 10% water samples. DNA isolated from positive cultures was used to perform spoligotyping and mycobacterial interspersed repetitive units-variable number of tandem repeats (MIRU-VNTR) analysis. Typing of environmental and clinical isolates revealed partial overlap in *Mtb* diversity present in the area^34^. These findings provided the first molecular evidence of detecting *Mtb* in environment. However, the screening strategy employed was expensive, time consuming, labor intensive, and required culturing hundreds of samples. Additionally, spoligotyping and MIRU-VNTR do not provide information on drug resistance and may lack sufficient data required to study transmission networks. Our novel sampling method allowed detection of *Mtb* on surfaces by direct swab sampling. By pooling samples, we further sequenced *Mtb* from environment samples for lineage identification, phylogenetic analysis, and variant calling. To the best of our knowledge, this is the first study to demonstrate the molecular detection of *Mtb* in environment using targeted qPCR assays and whole genome sequencing.

Previous studies have demonstrated that it is possible to detect *Mtb* from air samples in high-risk settings such as hospitals, clinics, and schools in TB endemic settings^35-37^. The utility of air sampling is that it can be used to directly characterize airborne exposure risk. However, it requires that the infectious individual have been present in the sampling location within minutes of the sampling, which limits its sensitivity in communal settings. Additionally, air sampling requires more sophisticated equipment and sampling that is conducted over a period of hours. Surface sampling, by contrast, can be performed in seconds using low-cost disposable swabs. It may be more sensitive, by detecting cumulative environmental contamination, but does not provide the temporal resolution for recent presence of an infectious individual as does air sampling. Surface sampling may therefore be more useful for identifying high risk locations than for identifying the presence of infectious individuals.

We detected *Mtb* DNA on multiple types of surfaces and locations, including vertical (walls) and horizontal ones (e.g. floors, bedding). While the study was not designed to provide comparative information about locations within rooms, we did not observe any systematic trends in positivity by location or material type. Given that *Mtb* travels in aerosolized droplet nuclei and can disseminate throughout rooms, it is likely to deposit on surfaces throughout rooms. Based on our experience collecting and analyzing samples, we speculate that surfaces that are less frequently cleaned (e.g. elevated or hard-to-reach surfaces) might be more likely to contain *Mtb* DNA. Additionally, personal utensils which frequently come in close proximity with the mouth of an infectious individual may also have higher chances of *Mtb* DNA detection if not cleaned thoroughly. Interestingly, we detected *Mtb* DNA on cigarette lighters from two cells analyzed on GeneXpert MTB/RIF Ultra. These lighters were re-swabbed after one week for hybrid capture assays, and we detected *Mtb* DNA on one of the lighters which matched with the *Mtb* genome from paired sputum culture. Additionally, a key variable which likely affects the recovery of *Mtb* DNA from surfaces is how frequently they are cleaned. Cells from the study prisons were regularly cleaned; by contrast, tuberculosis isolation rooms at the hospital only undergo light cleaning while occupied, followed by a terminal decontamination when the patient leaves. It’s possible that this (along with the high sputum smear grade) partially explains the higher recovery rate of *Mtb* from TB isolation rooms (supplementary material).

Using RNA hybrid capture assays, we were able to enrich and fully sequence *Mtb* genomes from ES swabs, which generated sufficient coverage for lineage identification and high-resolution variant calling. This is the first use of hybrid capture for enriching *Mtb* from non-sputum samples and demonstrates the potential for environmental sampling to inform our understanding of circulating *Mtb* diversity, including that of drug-resistance associated genotypes. ES swabs from two cells showed evidence of mixed infection and did not match with the paired sputum culture suggesting detection of an undiagnosed case or DNA from individual occupied the cell previously. We also detected *rpoB* (S450L) and 70 base pairs deletion in *katG* mutations conferring rifampin and isoniazid resistance respectively. However, no patient-derived clinical isolate containing rifampicin resistance was identified in that prison at the time of swab collection, despite a concomitant mass screening campaign in which >90% of prison residents participated. One possible explanation to these unexpected findings is frequent movement of incarcerated individuals between prisons cells. This observation is particularly important in case of drug resistant TB, where the infected individuals who usually share a cell with several healthy individuals continue to amplify the transmission as they move from one cell to another. This concerning finding suggests that ES sampling might provide information on *Mtb* strains which were missed through clinical screening.

The results of this study should be interpreted with the context of several limitations. First, we studied environments (hospital rooms, prisons) in which individuals with TB are present for prolonged periods of time, and positivity in environments where TB patients are present for shorter periods requires further investigation. Future studies should evaluate communal environments such as public transit, churches, schools, shops, bars and other places where individuals congregate. Second, we only sequenced ES swabs from a small subset of prison cells which had high ES swab positivity. Therefore, our study lacks data to interpret how frequently the *Mtb* DNA detected on the surface was indeed from the active patient present in that environment or we detected *Mtb* DNA accumulated over the period in the same prison cells. Lastly, our study lacks data on *Mtb* culture from environment samples. We attempted to culture *Mtb* from 75 swabs collected from positive prison cells (data not shown). However, none of the swab cultures were positive for *Mtb* after several weeks of incubation. This could be due to differentially culturable organisms or *in vitro* competition with other environmental organisms.

## 5. Conclusions

In summary, we found that *Mtb* DNA is frequently detectable in environments recently occupied by individuals with active TB. We demonstrated that environmental sampling for *Mtb* can be performed using a simple swabbing procedure and assayed on an automated molecular platform widely used in low- and middle-income countries. We further found that complete *Mtb* genome sequences can be recovered from environmental samples using an RNA-bait hybrid capture approach, with sufficient depth to allow variant calling and phylogenetic analysis. Our findings highlight the potential for harnessing information derived from environmental sampling in studying TB transmission.

## Data Availability

Data supporting the findings of this manuscript are available in the Supplementary Information files or from the corresponding author upon request. Access to the supplementary figure 2 is available from the corresponding author upon request Our bioinformatic pipeline is available at https://github.com/ksw9/mtb_pipeline.

## Data availability

Data supporting the findings of this manuscript are available in the Supplementary Information files or from the corresponding author upon request. Access to the **supplementary figure 2** is available from the corresponding author upon request Our bioinformatic pipeline is available at https://github.com/ksw9/mtb_pipeline.

## CRediT authorship contribution statement

**Renu Verma**: Study design; method development, validation, and data collection; Formal analysis, Visualization; Writing – original draft & editing. **Flora Martinez Figueira Moreira**: Sample collection; data collection; Writing – review & editing. **Agne Oliveira do Prado Morais**: Sample collection; Writing – review & editing. **Katharine S. Walter:** Writing – original draft & editing, data analysis and visualization. **Paulo César Pereira dos Santos:** Sample collection; data collection; Writing – review & editing. **Eugene Kim**: Data collection; Writing – review & editing. **Thiego Ramon Soares:** Sample collection; Writing – review & editing. **Rafaele Carla Pivetta de Araujo:** Sample collection; Writing – review & editing. **Bruna Oliveira da Silva**: Sample collection; Writing – review & editing. **Andrea da Silva Santos:** Sample collection; Writing – review & editing. **Julio Croda:** Study design; Supervision; Formal analysis; Writing – review & editing **Jason R. Andrews**: Conceptualization; Supervision; Study design; Writing – original draft & editing; Funding acquisition; Formal analysis; sample collection.

## Acknowledgements

We thank the prison staff for their support and incarcerated individuals from prisons Penitenciária Estadual de Dourados (PED) and Estabelecimento Penal Jair Ferreira de Carvalho (EPJFC), Brazil for collecting ES swabs from their cells. We also thank Leonardo Martinez from Boston University for his support.

---

**Supplementary Figure 1.**
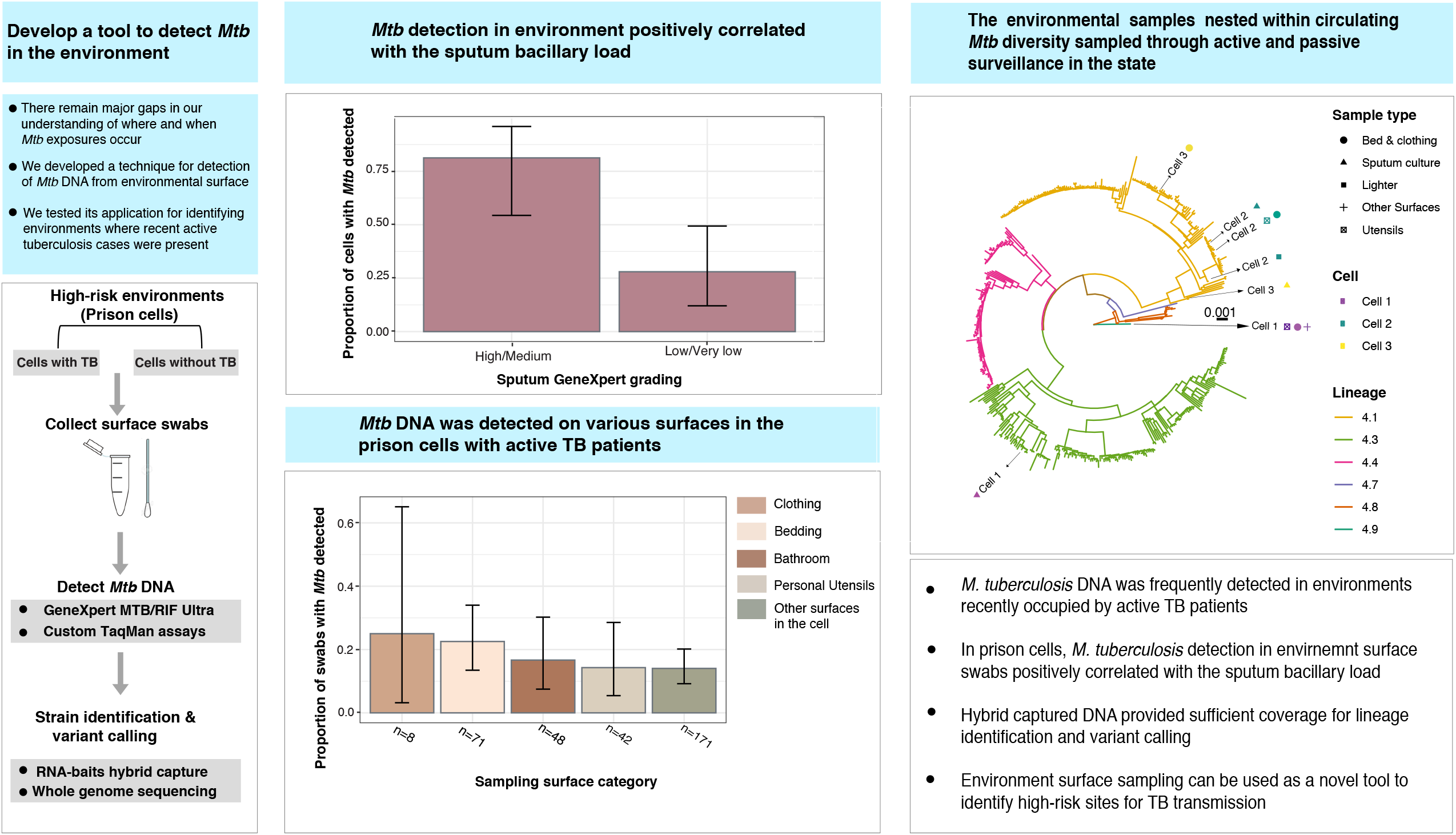
RD-9 TaqMan assay standard curve for *Mtb* genome copies quantification. The assay was performed in duplicates.

**Supplementary Figure 2.**
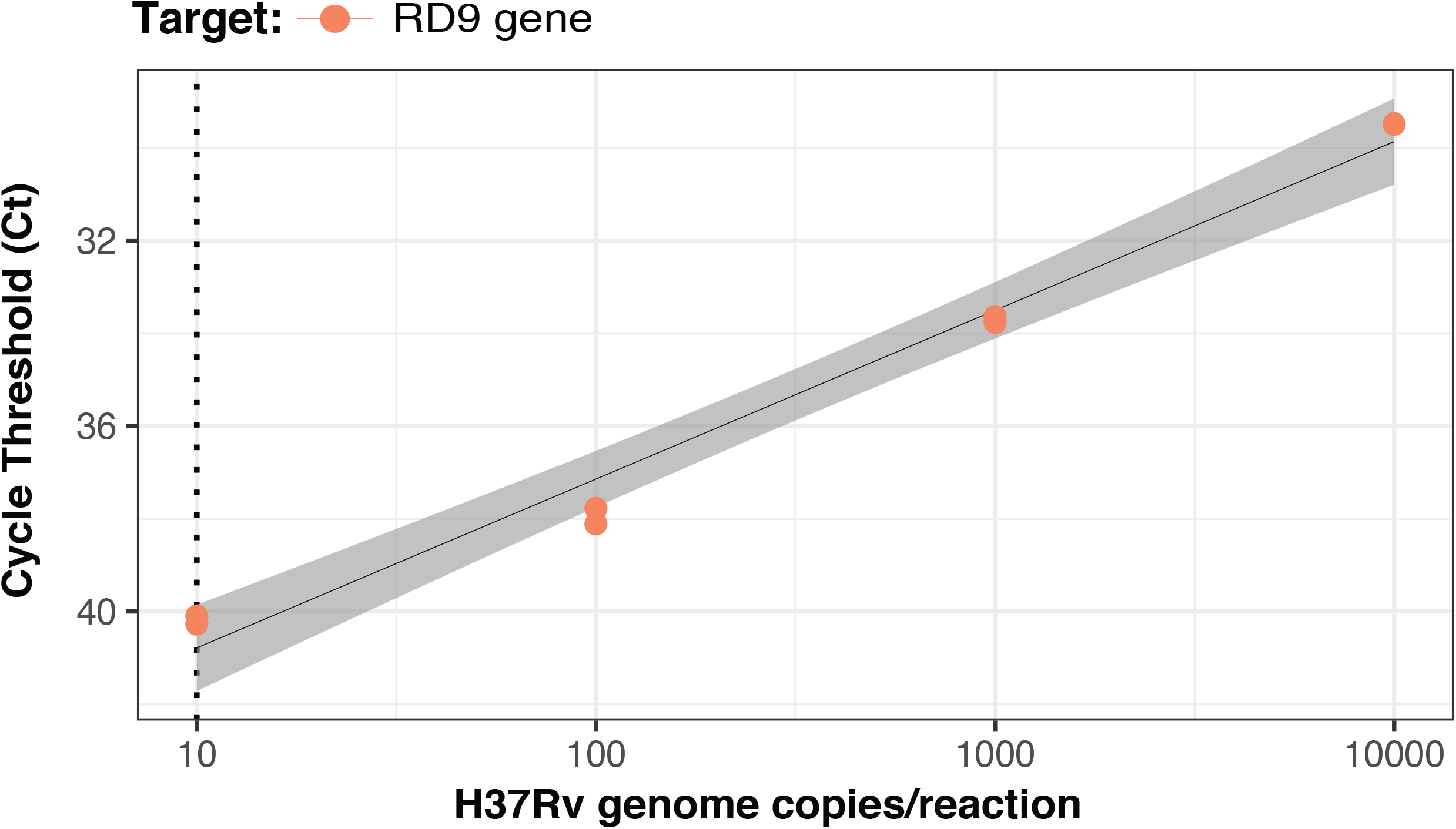
**(A)** A nurse at the Federal University of Mato Grosso do Sul, preparing to swab for the tuberculosis bacterium at the Penitenciária Estadual de Dourados, Brazil. **(B)** A study staff member teaching the incarcerated individuals how to collect environment surface swabs.

**Supplementary Figure 3.**
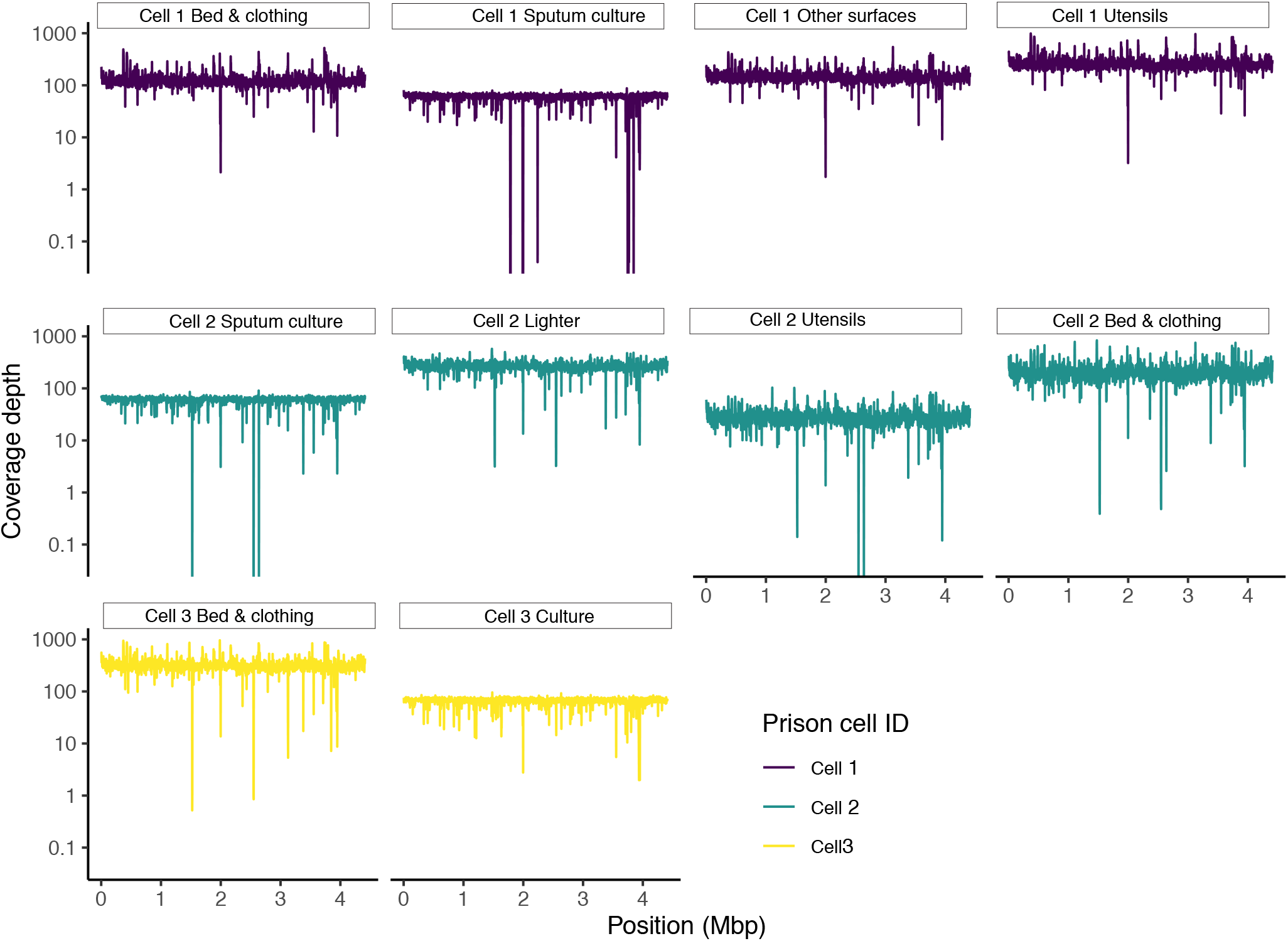
Coverage depth of paired environmental and culture samples. Coverage of the H37Rv reference genome determined with *mosdepth*. The seven captured ES swab samples had a mean genomic coverage of 137X (std. dev, 39) and median coverage of 135X. A mean of 98% of the *Mtb* genome had a depth of at least 10X; 65% had a depth of at least 100X.

