## Supplemental material for "Detection of *M. tuberculosis* in the environment as a tool for identifying high-risk locations for tuberculosis transmission"

**METHODS**

**Assay development and validation**

We first performed a pilot study at Stanford Hospital, CA, USA, to develop and validate the environment sampling assays. We collected a total 96 ES swabs from three, negative-pressure TB isolation rooms. Each room was occupied with one active, culture-confirmed pulmonary TB patient, and the surfaces were swabbed within an hour after the patient was discharged, before room decontamination. We collected ES swabs from various surfaces in the room which included beds, eating trays, bathrooms, floors, walls, and windows. A complete list of surfaces which were swabbed for *Mtb* DNA detection is provided in **Supplementary Table 1**.

Sterile Hydraflock swabs (Longhorn Primeswab LH-11-10) were used to collect ES swabs. Surfaces covering an area roughly the size of an adult palm (approx. 70 cm^2^) were swabbed with a hyraflock swab that was pre-wet with buffer (1X TE or 1X PBS). After collection, the ES swabs were immediately dispensed in 500ul buffer and transported to the molecular lab. We tested the *Mtb* DNA detection methods on two different platforms: GeneXpert MTB/RIF Ultra (Cepheid, Sunnyvale, USA) and StepOnePlus real time PCR machine (Applied Biosystems) using insertion sequence (IS) 6110 and the region of difference 9 (RD-9) custom TaqMan assays described previsouly. IS6110 is typically present in multiple, but variable numbers of copies in the *Mtb* genome, making it a sensitive target for detection but precluding quantification of genome copies, whereas RD-9 is present as a single copy, enabling genome copy quantification.

Seventy-six swabs from three TB isolation rooms were analyzed on custom TaqMan assays*.* Briefly, we collected ES swabs in 500ul 1X TE buffer (pH=7.4) (Millipore Sigma) for DNA extraction. DNA was extracted using QIAamp DNA mini kit (Cat # 51304). All qPCR reactions were performed in 20ul volume using TaqMan™ Environmental Master Mix 2.0 (Cat# 4396838) with 500nM of each forward and reverse primers and 250nM of probes. The samples were analyzed in technical duplicates on a StepOnePlus Real Time PCR using the following cycling conditions; initial denaturation (95ºC;10 minutes), 40 cycles (denaturation 95 ºC 15 seconds and 60ºC 30 seconds annealing and extension). The samples were considered positive only when the amplification was observed in both duplicates of IS6110 and RD-9 assays. DNA copies were quantified using a RD-9 standard curve derived from *Mtb* H37Rv ATCC strain (Catalogue#25618DQ) (**Supplementary Figure 1**).

We then compared IS6110 and RD-9 assays with GeneXpert MTB/RIF Ultra (Cepheid, Sunnyvale, CA) on additional 20 ES swabs to simultaneously to compare the sensitivities of two assays. For GeneXpert analysis, samples were collected in 500ul sterile 1X PBS (pH=7.4) (Cytiva HyClone™) and used directly without manual DNA extraction. Briefly, the swabs were vortexed for 30 seconds and the samples were mixed with Xpert MTB/RIF buffer in 1:1 (sample to buffer ratio). The samples were vortexed for 5 seconds followed by 5 minutes incubation at room temperature. The entire 1ml sample was loaded into GeneXpert MTB/RIF Ultra cartridge for *Mtb* DNA detection. GeneXpert *Mtb* load was graded semiquantitatively as high (Ct <16), medium (Ct 16<22), low (Ct 22–28), very low (Ct >28) and trace).

**RESULTS**

***Mtb* DNA detection in TB isolation rooms**

For assay development and validation, we collected ES swabs from three negative pressure TB isolation rooms at the time of patient discharge. The three culture-confirmed pulmonary tuberculosis patients had maximum AFB (Acid-Fast Bacillus) smear results of 4+, 3+ and negative. *Mtb* DNA was detected in 19/76 (25%) of the ES swabs analyzed on IS6110 and RD-9 custom assays. ES swabs *Mtb* positivity by Taqman real-time PCR was 10/20 (50%) in the room of the participant with 4+ AFB, 9/23 (39%) in room of the participant with 3+ AFB, and all were negative (0/33) in the room of the smear-negative participant. We did not detect any *Mtb* DNA in 30 control ES swabs contemporaneously collected from lab and office spaces.

We then compared detection of *Mtb* DNA from paired ES swabs assayed on GeneXpert MTB/RIF Ultra and TaqMan IS6110 and RD-9 custom qPCR assays. Among 20 paired samples, eight pairs were positive and 6 were negative for *Mtb* DNA on both GeneXpert and TaqMan IS6110 and RD-9 assays. Four swabs were positive only on GeneXpert and two only on TaqMan assays (**Supplementary Table 1**).
